# Early nimodipine treatment in reversible cerebral vasoconstriction syndrome: a serial transcranial Doppler study

**DOI:** 10.1101/2024.06.07.24308638

**Authors:** Soohyun Cho, Mi Ji Lee

## Abstract

**Background:** Although nimodipine is commonly used to treat reversible cerebral vasoconstriction syndrome (RCVS), its disease-modifying effects are not yet understood. We aimed to investigate the effect of nimodipine and treatment initiation timing on the prevention of worsened vasoconstriction.

**Methods:** We prospectively recruited patients with recent-onset (within 3 weeks of the first thunderclap headache), angiogram-proven RCVS. All participants underwent transcranial Doppler (TCD) examination to evaluate the mean flow velocities (MFVs) of the bilateral middle cerebral arteries at baseline and were serially followed-up after 10, 20, 30, and 90 days. Oral nimodipine treatment was initiated after the baseline TCD study, and patients were split into early and late treatment groups based on the time from onset to initial nimodipine administration. To estimate the worsening of vasoconstriction, we calculated the area of total time spent with MFVs above the baseline value (“MFV area”). We tested the correlation between the number of days from onset to treatment initiation and MFV area and performed linear regression analysis to examine the independent association between earlier treatment and MFV area.

**Results:** A total of 32 patients with RCVS (mean age: 51.5 ± 10.3 years; 91% female) completed this study. Baseline TCD assessment was performed at a mean of 7.1 ± 4.2 days after thunderclap headache onset. We observed that earlier treatment with nimodipine correlated with reduced MFV area (r = 0.37, *p* = 0.038). Furthermore, this association remained significant after adjusting for other clinical variables (regression coefficient 0.673, adjusted *p* = 0.023) in the multivariable linear regression.

**Conclusions:** Early oral nimodipine treatment prevents worsening of vasoconstriction, suggesting that nimodipine may have a disease-modifying effect in RCVS treatment.

## Introduction

Reversible cerebral vasoconstriction syndrome (RCVS) is characterized by thunderclap headaches (TCHs) with vasospasms of the cerebral arteries, which may lead to neurological complications such as stroke and subarachnoid hemorrhage.^1,2^ Angiographic findings in RCVS exhibit dynamic changes over time; vasoconstriction progressively worsens and regresses spontaneously, with maximal involvement approximately 3 weeks after onset.^3,4^

Currently, there are no evidence-based guidelines for the treatment of RCVS. Nimodipine is frequently administered for RCVS because it blocks the vasoconstrictive cascade.^2,5,6^ The effect can be immediately observed during intra-arterial infusion and even intravenous treatment.^7–9^ In addition, we previously reported that nimodipine shortens the clinical course of RCVS, as defined by the duration of TCH recurrence.^10^ However, the disease-modifying effect of oral nimodipine, i.e., preventing the temporal worsening of cerebral hemodynamics, is not well understood.^3,4^ This may be attributed to a lack of placebo-controlled trials and the complexity of natural hemodynamic changes in RCVS.

Therefore, we aimed to document the restorative effect of nimodipine on cerebral vasoconstriction by considering the timing of nimodipine treatment. Specifically, we hypothesized that earlier nimodipine administration could be linked to better outcomes by preventing worsening of and repairing cerebral vasoconstriction in RCVS. In this study, we prospectively evaluated the cerebral blood flow for 3 months following oral nimodipine treatment in patients with RCVS to investigate the effect of the drug on cerebral hemodynamics.

## Methods

Our article was reported according to the STROBE guideline.

### Patients and Study Setting

We prospectively recruited patients with angiogram-proven RCVS who visited the Samsung Medical Center, Seoul, South Korea, between June 2019 and November 2021. We included patients who (1) clearly remembered the mode of onset of TCH, (2) reported the time from headache onset to its peak as < 1 min, and (3) had the first TCH within the last 3 weeks. The 3-week criterion was used to ensure a linear association between the time from onset to treatment and the initial severity of vasoconstriction. All patients were interviewed by two experienced neurologists specializing in headache disorders (M.J.L. and S.C.) and were primarily evaluated using brain imaging to confirm the diagnosis of RCVS.

All patients underwent brain MRI and magnetic resonance angiography (MRA) for the differential diagnosis following inclusion. Thereafter, we evaluated the mean flow velocities (MFVs) of the bilateral middle cerebral arteries (MCAs) using transcranial Doppler (TCD) and conducted serial follow-ups at 10, 20, 30, and 90 days from the initial assessment (Figure S1). To determine the extent of the worsening of vasoconstriction, we calculated the “MFV area,” which was defined as the total time that the MFV was above the baseline MFV (Figure 2). As all patients exhibited vasoconstriction at baseline and the MFV measured at the initial visit did not reflect their actual baseline hemodynamics when healthy, we set the baseline MFV using the follow-up TCD study at 90 days, when vasoconstriction was normalized. The normalization of vasoconstriction was ensured by performing follow-up MRA at this time point. Oral nimodipine treatment was initiated immediately after the initial TCD study at 30– 60 mg every 8–12 hours per day and was maintained until the end of the study (90 days).

We hypothesized that nimodipine would prevent the worsening of vasoconstriction, which typically occurs in the first 3 weeks after onset. However, as nimodipine is prescribed to all patients at our center, we were unable to compare its effect with those of non-use. Instead, we investigated the effect of the timing of initial nimodipine administration on the clinical course of vasoconstriction. During the natural course of RCVS, MFV progressively increases during the first few weeks and peaks approximately 3 weeks after onset (Figure 1A). We hypothesized that nimodipine would attenuate the worsening of vasoconstriction by reducing MFV area (Figure 1B). However, if nimodipine treatment was initiated later, we speculated that the maximal MFV would be greater than that in the early treatment scenario but normalize faster than that in the natural course (Figure 1C).

**Figure 1.**
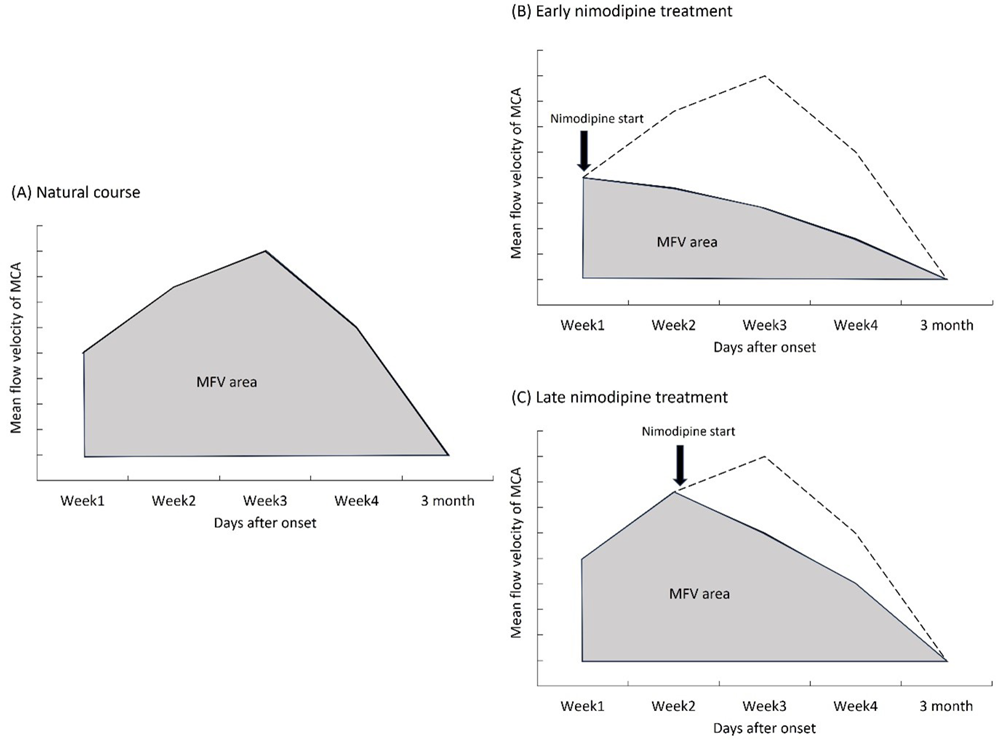
Hypothetical plots of the temporal course of the MCA mean flow velocity (MFV) and MFV area in reversible cerebral vasoconstriction syndrome in different treatment scenarios. (A) Natural course, (B) early nimodipine treatment, and (C) late nimodipine treatment. Dashed line represents natural course. MCA, middle cerebral artery

### Statistical Analyses

Descriptive data are presented as the number (percentage) or mean ± SD. We used a linear mixed model with random effects (patients) to assess the change in MFV of the MCA from baseline to follow-up. Using the difference in MFV from baseline during follow-up, we constructed a scatterplot using a locally weighted scatterplot smoothing (LOESS) curve. In addition, we analyzed the individual changes in the MCA MFV for each patient. We defined a deterioration in MFV as > 20 cm/sec greater than the baseline value.^11^ We used the MFV area to test whether earlier administration of nimodipine affected the degree of increased vasoconstriction. Pearson’s correlations between the MFV areas and clinical variables, including the number of days from onset to treatment initiation, were performed. Subsequently, we conducted stepwise multiple linear regression analysis to identify the clinical variables associated with MFV areas. The MFV area was the dependent variable, whereas the candidate independent variables included days from onset to treatment initiation, sex, age, body mass index (BMI), TCH severity, number of TCHs, presence of trigger factors, and comorbid migraine. Additionally, we used Fisher’s exact test or the Mann–Whitney U test to compare categorical and continuous variables between early nimodipine treatment (< 7 days after onset) and late nimodipine treatment (≥ 7 days after onset). All statistical analyses were conducted using SPSS software (version 22.0; IBM Corporation, Armonk, NY, USA). Statistical significance was set at *p* < 0.05 (two-tailed).

### Standard Protocol Approvals, Registrations, and Patient Consents

The Institutional Review Board of Samsung Medical Center approved this study (IRB no. 2019-03-104-001). Written informed consent was obtained from all patients prior to recruitment.

### Data Availability

Anonymized data not published within this article will be made available by request from any qualified investigator.

## Results

### Patient Characteristics

We screened 47 patients with TCH, and 36 patients were diagnosed with angiogram-proven RCVS. After excluding three patients who had their first TCH after three weeks from onset, 33 patients were enrolled for the study. One patient did not complete serial TCDs. Finally, a total of 32 patients were analyzed in this study. Table 1 presents the demographic and clinical characteristics of the individuals at baseline. Patients had a mean age of 51.5 ± 10.3 years (range, 29–73 years) and 90.6% were female. The mean number of TCHs at baseline was 2.7. The mean days from onset to initiation of nimodipine was 7.1 ± 4.2 (range from 0–15 days). For the early treatment group (n = 15), the time was 3.5 ± 1.7 days, while for the late treatment group (n = 17), it was 10.2 ± 2.9 days (p < 0.001).No significant differences in demographic or clinical characteristics were observed between the groups (Table 1).

**Table 1.**
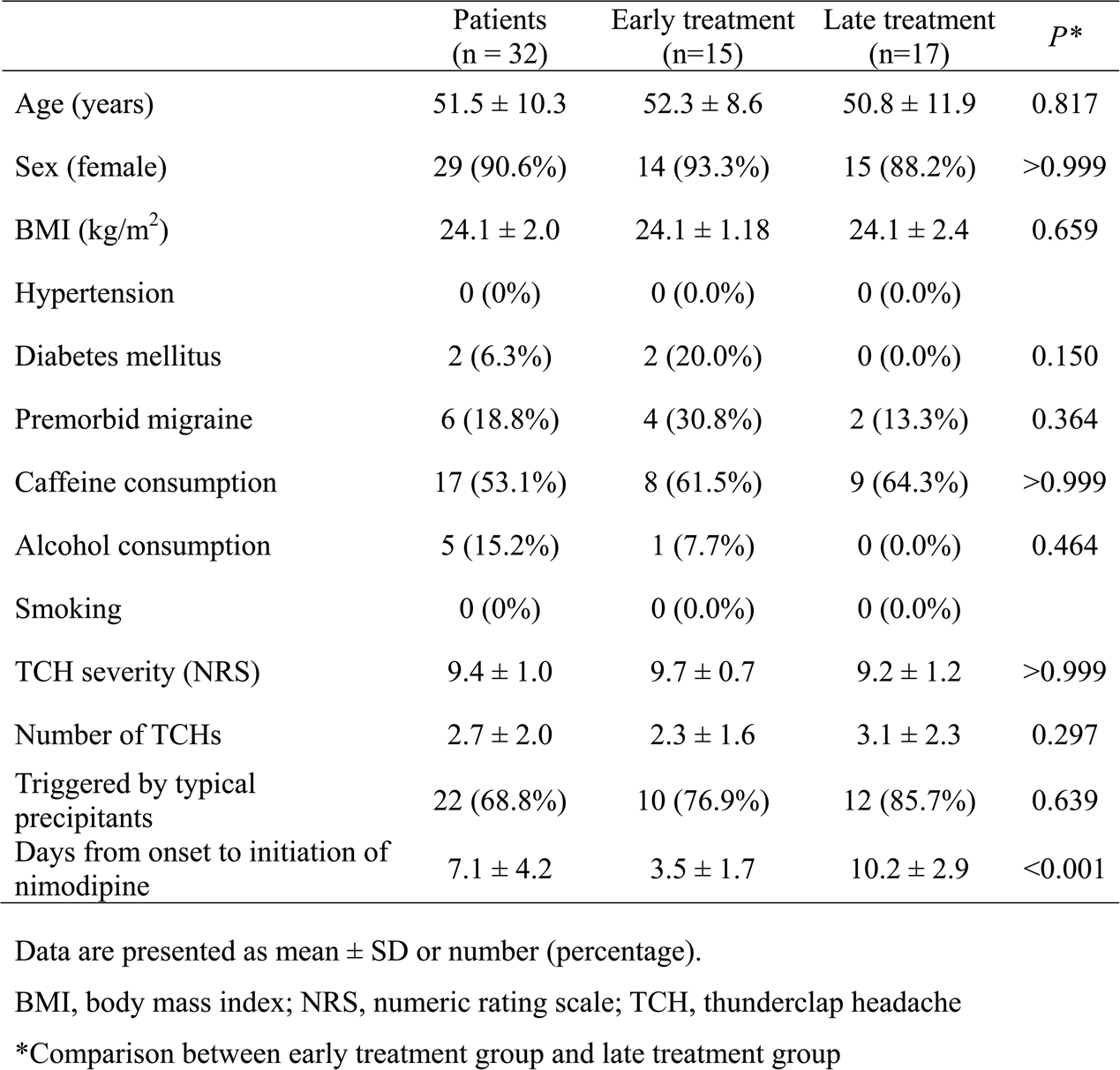
Demographic and clinical characteristics of patients at baseline visit.

|  | Patients<br>(n = 32) | Early treatment<br>(n=15) | Late treatment<br>(n=17) | *P* * |
| --- | --- | --- | --- | --- |
| Age (years) | 51.5 ± 10.3 | 52.3 ± 8.6 | 50.8 ± 11.9 | 0.817 |
| Sex (female) | 29 (90.6%) | 14 (93.3%) | 15 (88.2%) | >0.999 |
| BMI (kg/m<sup>2</sup>) | 24.1 ± 2.0 | 24.1 ± 1.18 | 24.1 ± 2.4 | 0.659 |
| Hypertension | 0 (0%) | 0 (0.0%) | 0 (0.0%) |  |
| Diabetes mellitus | 2 (6.3%) | 2 (20.0%) | 0 (0.0%) | 0.150 |
| Premorbid migraine | 6 (18.8%) | 4 (30.8%) | 2 (13.3%) | 0.364 |
| Caffeine consumption | 17 (53.1%) | 8 (61.5%) | 9 (64.3%) | >0.999 |
| Alcohol consumption | 5 (15.2%) | 1 (7.7%) | 0 (0.0%) | 0.464 |
| Smoking | 0 (0%) | 0 (0.0%) | 0 (0.0%) |  |
| TCH severity (NRS) | 9.4 ± 1.0 | 9.7 ± 0.7 | 9.2 ± 1.2 | >0.999 |
| Number of TCHs | 2.7 ± 2.0 | 2.3 ± 1.6 | 3.1 ± 2.3 | 0.297 |
| Triggered by typical precipitants | 22 (68.8%) | 10 (76.9%) | 12 (85.7%) | 0.639 |
| Days from onset to initiation of nimodipine | 7.1 ± 4.2 | 3.5 ± 1.7 | 10.2 ± 2.9 | <0.001 |

Data are presented as mean ± SD or number (percentage).
BMI, body mass index; NRS, numeric rating scale; TCH, thunderclap headache
\*Comparison between early treatment group and late treatment group

### MFV and MFV Area

All the patients received nimodipine treatment and completed five serial TCD studies. The initial TCD assessment and subsequent immediate treatment initiation were conducted at a mean interval of 7.1 ± 4.2 days after onset. The mean follow-up duration was 111 ± 11.3 days.

The linear mixed model analysis revealed that the MFV of the MCA gradually decreased with time after nimodipine treatment (*p* < 0.001) as follows: 81.9 ± 23.2 cm/sec, 77.3 ± 28.9 cm/sec, 74.4 ± 23.9 cm/sec, 75.6 ± 23.3 cm/sec, and 69.3 ± 18.7 cm/sec at the initial visit and the 10-, 20-, 30-, and 90-day follow-ups, respectively (Figure 2). When patients were individually analyzed, all except two (93.8%, n = 30/32) showed no deterioration in MFV from baseline after treatment.

**Figure 2.**
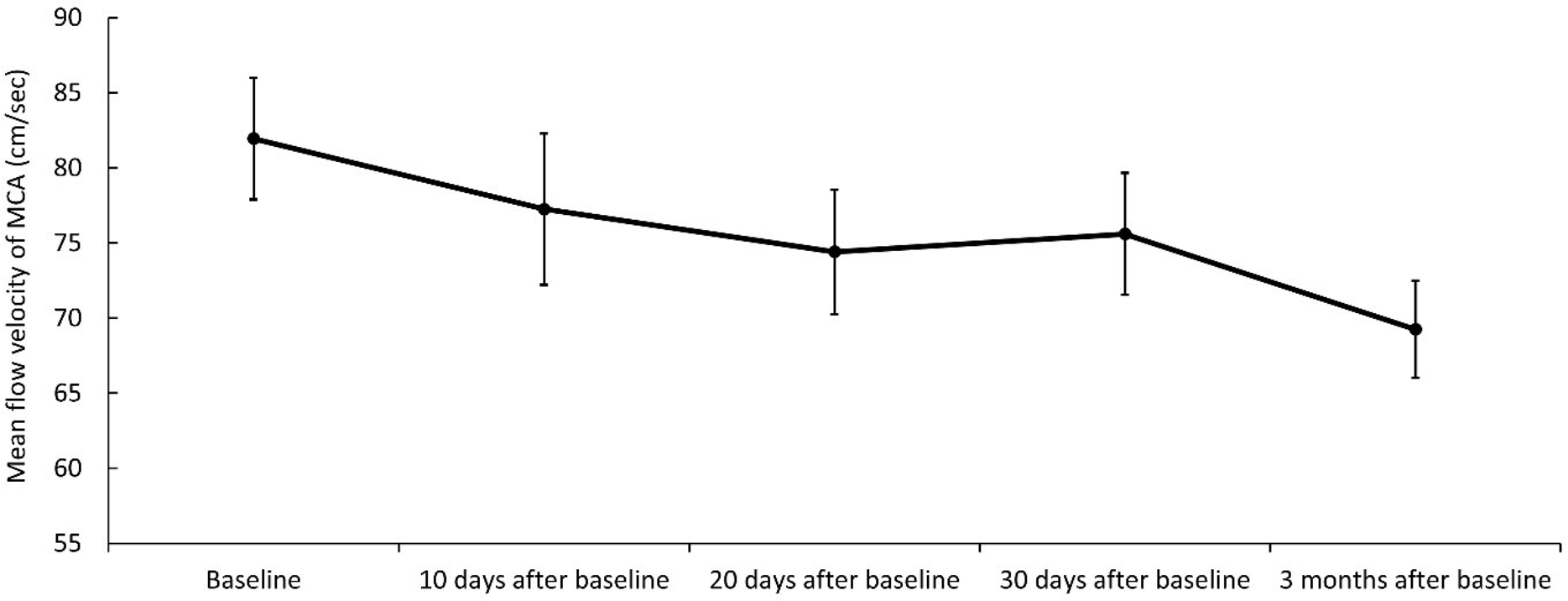
Temporal course of the MFV of the middle cerebral artery from baseline to final follow-up. Line indicates MFV of the middle cerebral artery over time. Error bars indicate SEM. MFV, mean flow velocity

The correlations between the MFV area and number of days from onset to initial nimodipine administration are shown in Figure 3. Earlier treatment correlated with lower MFV area (r = 0.37, *p* = 0.038).

**Figure 3.**
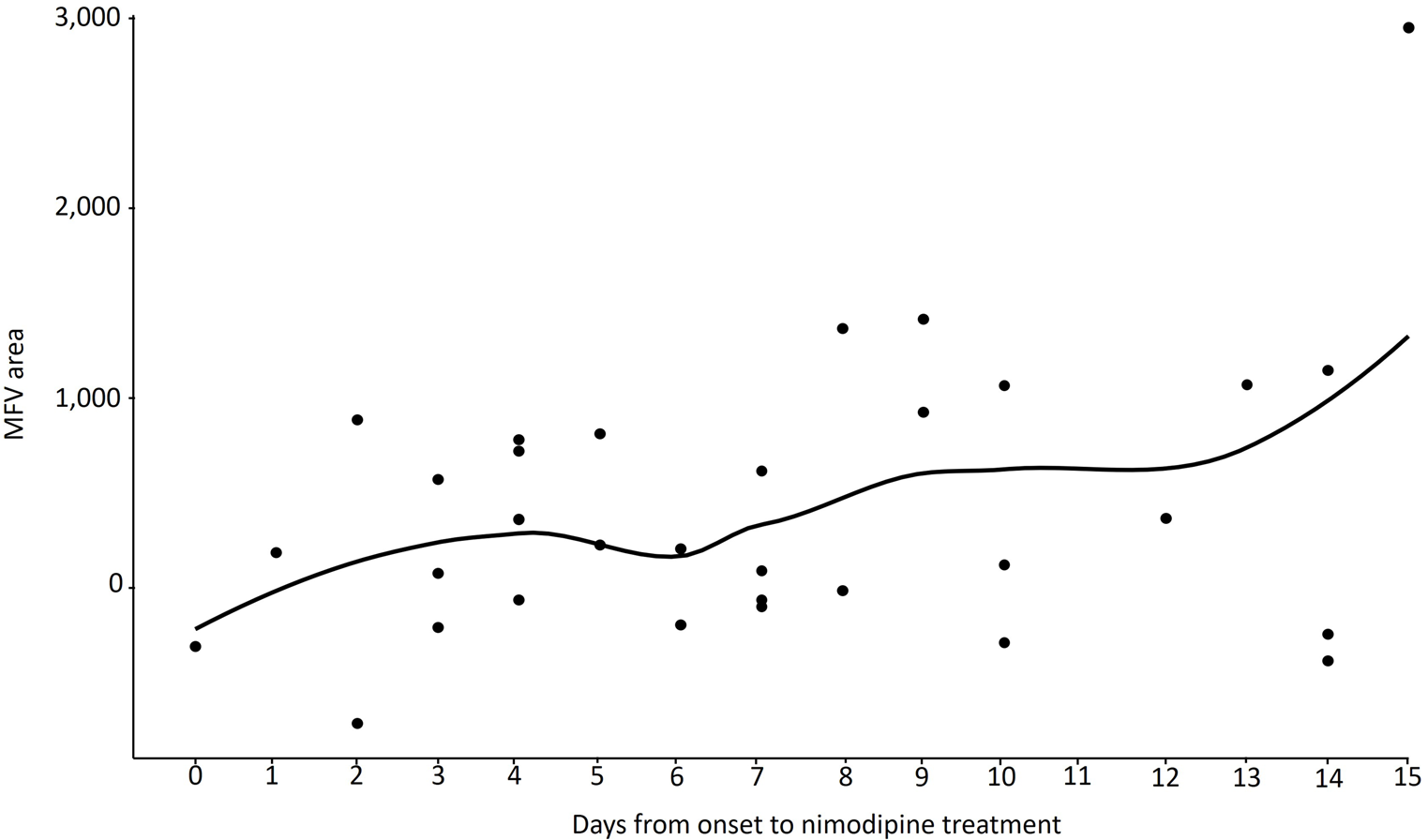
Correlation between MFV area and days from onset to nimodipine treatment initiation. Dots represent individual values. Line represents locally weighted scatterplot smoothing regression curve. MFV, mean flow velocity

### MFV and MFV Area in Early and Late Treatment Groups

The MFV gradually decreased in both the early (n = 15) and late treatment groups (n = 17) (Figure 4). MFVs in the early treatment group were lower than the untreated baseline MFVs of the late treatment group at the same time point after onset (69.8 ± 20.1 vs. 86.1 ± 24.4 cm/sec, *p* = 0.032; Figure 4). No differences were observed between the two groups for the other follow-up periods. Additionally, the MFV area was lower in the early treatment group than the late treatment group, although this difference was not significant (221.3 ± 471.5 vs. 589.7 ± 864.0, *p* = 0.278).

**Figure 4.**
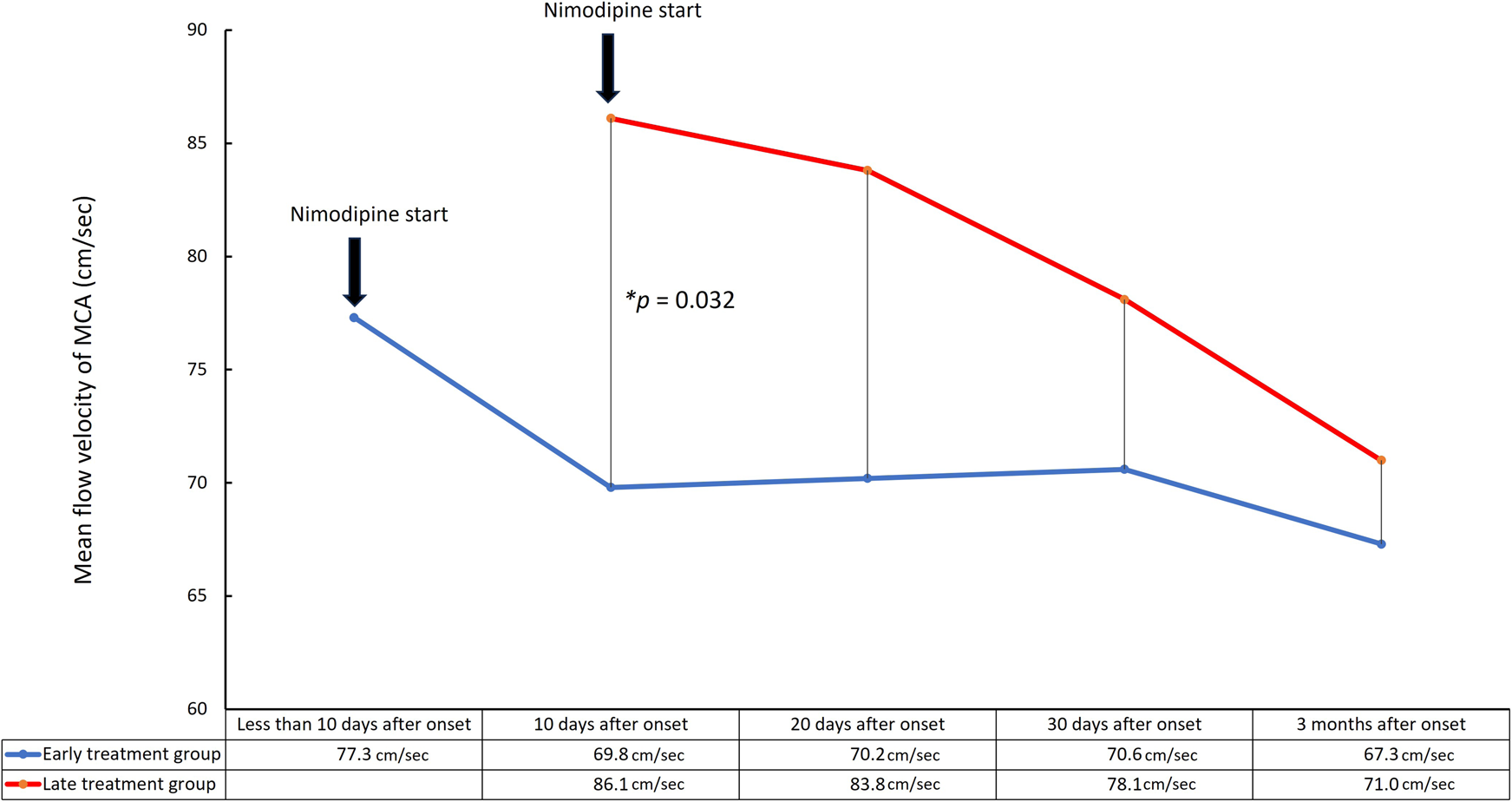
Comparison of the temporal course of the MCA mean flow velocity (MFV) and MFV area according to treatment timing. Blue line indicates MFV of the MCA of the early treatment group (< 7 days between onset and nimodipine initiation) and red line indicates that of the late treatment group (≥ 7 days between onset and nimodipine initiation). Asterisk indicates significant difference between MFV of early treatment group and untreated baseline MFV of late treatment group (*p* = 0.032). MCA, middle cerebral artery

### Determinants of MFV Area

To assess the independent effect of the timing of nimodipine treatment on the clinical course of cerebral hemodynamics, we conducted a stepwise multivariable linear regression analysis of the MFV area with other clinical covariates (Table 2). During the analysis, covariates such as sex, age, BMI, TCH severity, number of TCHs, presence of trigger factors, and comorbid migraine did not remain in the final model. In the final model, treatment timing was independently associated with MFV area (model 1: standardized coefficient ß = 0.673, *p* = 0.023; model 2: ß = 0.734, *p* = 0.006 and model 2: ß = 0.492, *p* = 0.038 after controlling for the effect of BMI).

**Table 2.** Multivariable linear regression analysis of nimodipine administration timing and worsening of vasoconstriction (measured using mean flow velocity area.

| | $R^2$ | Adjusted $R^2$ | Standardized<br>coefficient $\beta$ | $p$-value | t-value |
| --- | --- | --- | --- | --- | --- |
| Model 1 | 0.673 | 0.453 |  |  |  |
| Days after onset |  |  | 0.673 | 0.023 | 2.732 |
| Model 2 | 0.831 | 0.691 |  |  |  |
| Days after onset |  |  | 0.734 | 0.006 | 3.710 |
| BMI |  |  | 0.492 | 0.038 | 2.483 |

BMI, body mass index

## Discussion

In this study, oral nimodipine treatment was effective in preventing the worsening of vasoconstriction in RCVS, as evidenced by the following: (1) during the nimodipine treatment, MFVs of MCA gradually decreased in almost all patients, with more than 90% of patients exhibiting no significant increase in MFV from baseline; (2) MFVs in the early treatment group were lower than the untreated baseline MFVs of the late treatment group at the same time point after onset; and (3) the timing of nimodipine treatment was independently associated with MFV area, indicating that earlier nimodipine treatment administration was correlated with an improved temporal course of vasoconstriction.

Our first finding was that nimodipine treatment led to a decrease in the MFVs for almost all patients. Approximately 93% of patients showed no deterioration in the MFV of the MCA after treatment. In RCVS, vasoconstriction typically worsens over time and peaks at approximately three weeks from onset.^3,4^ Centripetal propagation, a distinctive phenomenon characterized by the propagation of vasoconstriction from distal arterioles to proximal medium-sized arteries, was rarely observed in our study, whereas approximately half of patients with RCVS showed centripetal propagation in a previous study.^12^ This finding may support the role of nimodipine in preventing exacerbated vasoconstriction.

The potential disease-modifying role of nimodipine is also illustrated by the significant decrease in MVF by early nimodipine treatment compared to that of the late treatment group at the same time point. This finding indicates that early nimodipine administration may alter the natural disease course in RCVS. Given the lack of randomized, placebo-controlled studies on nimodipine, its disease-modifying role has not been investigated. Although we did not implement a placebo arm, our study setting enabled us to compare nimodipine-treated and untreated groups by recruiting patients at different time points after onset. Another strength of our study is that nimodipine was initiated within 7 days after onset in the early treatment group, which is much earlier than the time to treatment reported in previous studies.^4,13,14^

In our study, the timing of oral nimodipine treatment was independently associated with the MFV area, indicating that earlier nimodipine treatment leads to a better temporal course of vasoconstriction. The efficacy of nimodipine in mitigating vasoconstriction in RCVS can be observed immediately after intra-arterial injection.^7,15^ However, the effect of oral nimodipine on vasoconstriction in patients with RCVS was previously unclear. Our findings suggest that early oral administration may interrupt the temporal progression of vasoconstriction in RCVS. It is known that nimodipine effectively inhibits transmembrane Ca^2+^ influx following the depolarization of smooth muscle cells, thereby reducing Ca^2+^-dependent activation of the contractile machinery.^16^ Recent preclinical data have identified additional mechanisms by which nimodipine may prevent detrimental vascular responses to spreading depolarization in the ischemic brain.^17^ Additionally, nimodipine can restore impaired neurovascular coupling, preventing transient and inappropriate constriction of small arteries and arterioles.^18^ Based on these observations, nimodipine may exert a disease-modifying effect during RCVS treatment. In this prospective cohort study, we performed serial follow-up assessments at various time points, combined with the evaluation of clinical variables, which provided a more thorough understanding of the disease course and the impact of nimodipine treatment in RCVS than previous studies. Nevertheless, our study has several limitations. First, we could not control the timing of nimodipine treatment because of the severity of TCHs and the scheduling of TCD examinations; thus, the treatment initiation time varied among patients. However, the distribution of patients according to treatment timing in 3 weeks of treatment initiation period allowed for meaningful comparisons. Second, the study did not include a control group for comparison owing to ethical concerns, making it challenging to draw definitive conclusions regarding the efficacy of nimodipine in RCVS treatment. Third, our study was conducted at a single medical center, which may limit the generalizability of the results. However, in a recent survey, we found similar patient characteristics among headache clinics in South Korea^19^; therefore, we anticipate that the effects of nimodipine are unlikely to differ by institution. Forth, we did not observe a correlation between improvement in the MFV or MFV area after nimodipine treatment and clinical outcomes. However, because we previously found an effect of nimodipine on clinical outcomes^10^, future larger studies may identify a such a correlation. Finally, the sensitivity of TCD may not be optimal in detecting centripetal propagation. Thus, our finding of the role of nimodipine in preventive centripetal propagation should be validated by using more sensitive imaging modalities.

## Conclusions

In conclusion, our study showed an independent association between the timing of oral nimodipine administration and exacerbation of cerebral vasoconstriction in patients with RCVS, demonstrating that early administration prevents the worsening of vasoconstriction. These results suggest that nimodipine exerts a disease-modifying effect in the treatment of RCVS.

## Acknowledgements

None

## Non-standard Abbreviations and Acronyms

BMI: body mass index
LOESS: locally weighted scatterplot smoothing
MCA: middle cerebral artery
MFV: mean flow velocity
NRS: numeric rating scale
RCVS: reversible cerebral vasoconstriction syndrome
TCD: transcranial Doppler
TCH: thunderclap headache

## References

1. Calabrese LH, Dodick DW, Schwedt TJ, Singhal AB. Narrative review: reversible cerebral vasoconstriction syndromes. Ann Intern Med. 2007;146:34–44.

2. Ducros A, Boukobza M, Porcher R, Sarov M, Valade D, Bousser M-G. The clinical and radiological spectrum of reversible cerebral vasoconstriction syndrome. A prospective series of 67 patients. Brain. 2007;130:3091–3101.

3. Chen SP, Fuh JL, Wang SJ, Chang FC, Lirng JF, Fang YC, Shia BC, Wu JC. Magnetic resonance angiography in reversible cerebral vasoconstriction syndromes. Ann Neurol. 2010;67:648–656.

4. Chen SP, Fuh JL, Chang FC, Lirng JF, Shia BC, Wang SJ. Transcranial color doppler study for reversible cerebral vasoconstriction syndromes. Ann Neurol. 2008;63:751–757.

5. Chen S-P, Fuh J-L, Lirng J-F, Chang F-C, Wang S-J. Recurrent primary thunderclap headache and benign CNS angiopathy: spectra of the same disorder? Neurology. 2006;67:2164–2169.

6. Singhal AB, Hajj-Ali RA, Topcuoglu MA, Fok J, Bena J, Yang D, Calabrese LH. Reversible cerebral vasoconstriction syndromes: analysis of 139 cases. Arch Neurol. 2011;68:1005–1012.

7. Linn J, Fesl G, Ottomeyer C, Straube A, Dichgans M, Bruckmann H, Pfefferkorn T. Intra-arterial application of nimodipine in reversible cerebral vasoconstriction syndrome: a diagnostic tool in select cases? Cephalalgia. 2011;31:1074–1081. doi: 10.1177/0333102410394673

8. Kass-Hout T, Kass-Hout O, Sun CH, Kass-Hout T, Ramakrishnan P, Nahab F, Nogueira R, Gupta R. A novel approach to diagnose reversible cerebral vasoconstriction syndrome: a case series. J Stroke Cerebrovasc Dis. 2015;24:e31–37. doi: 10.1016/j.jstrokecerebrovasdis.2014.08.023

9. Hockel K, Diedler J, Steiner J, Birkenhauer U, Ernemann U, Schuhmann MU. Effect of intra-arterial and intravenous nimodipine therapy of cerebral vasospasm after subarachnoid hemorrhage on cerebrovascular reactivity and oxygenation. World neurosurgery. 2017;101:372–378.

10. Cho S, Lee MJ, Chung C-S. Effect of nimodipine treatment on the clinical course of reversible cerebral vasoconstriction syndrome. Front Neurol. 2019;10:644.

11. Sharma S, Lubrica RJ, Song M, Vandse R, Boling W, Pillai P. The role of transcranial Doppler in cerebral vasospasm: a literature review. Subarachnoid Hemorrhage: Neurological Care and Protection. 2020:201–205.

12. Shimoda M, Oda S, Shigematsu H, Hoshikawa K, Imai M, Komatsu F, Hirayama A, Osada T. Clinical significance of centripetal propagation of vasoconstriction in patients with reversible cerebral vasoconstriction syndrome: a retrospective case-control study. Cephalalgia. 2018;38:1864–1875.

13. Hathidara M, Patel NH, Flores A, Cabrera Y, Cabrera F, Koch S. Transcranial Doppler findings in reversible cerebral vasoconstriction syndrome. J Neuroimaging. 2022;32:345–351.

14. Marsh EB, Ziai WC, Llinas RH. The need for a rational approach to vasoconstrictive syndromes: transcranial doppler and calcium channel blockade in reversible cerebral vasoconstriction syndrome. Case Rep Neurol. 2016;8:161–171.

15. Elstner M, Linn J, Müller-Schunk S, Straube A. Reversible Cerebral Vasoconstriction Syndrome: A Complicated Clinical Course Treated with Intra-Arterial Application of Nimodipine. Cephalalgia. 2009;29:677–682. doi: 10.1111/j.1468-2982.2008.01768.x

16. Scriabine A, Van den Kerckhoff W. Pharmacology of nimodipine. A review. Ann N Y Acad Sci. 1988;522:698–706.

17. Szabó Í, M. Tóth O, Török Z, Varga DP, Menyhárt Á, Frank R, Hantosi D, Hunya Á, Bari F, Horváth I. The impact of dihydropyridine derivatives on the cerebral blood flow response to somatosensory stimulation and spreading depolarization. Br J Pharmacol. 2019;176:1222–1234.

18. Dreier JP. The role of spreading depression, spreading depolarization and spreading ischemia in neurological disease. Nat Med. 2011;17:439–447.

19. Cho S, Kim B-K, Lee MJ. Clinical characteristics of reversible cerebral vasoconstriction syndrome: A large Korean multicenter study. Cephalalgia. 2023;43:288–288.

